# Time-to-event prediction in ALS using a landmark modeling approach, using the ALS Natural History Consortium Dataset

**DOI:** 10.1101/2024.11.15.24317346

**Authors:** David Schneck, Andres Arguedas, Annette Xenopoulos-Oddsson, Ximena Arcila-Londono, Christian Lunetta, James Wymer, Nicholas Olney, Kelly Gwathmey, Senda Ajroud-Driss, Ghazala Hayat, Terry Heiman-Patterson, Federica Cerri, Christina Fournier, Jonathan Glass, Alex Sherman, Mark Fiecas, David Walk

## Abstract

**Background and Objectives:** Times to clinically relevant events are a valuable outcome in observational and interventional studies, complementing linear outcomes such as functional rating scales and biomarkers. In ALS, there are several clinically relevant events. We developed dynamic prediction models for several of these times to events that can be used for clinical trial modeling and personal planning.

**Methods:** Landmark time-to-event analysis was implemented to determine the effect of patient characteristics on disease progression. Longitudinal data from 1557 participants in the ALS Natural History Consortium dataset were used. Five outcomes in the ALS disease progression were considered: loss of ambulation, loss of speech, gastrostomy, non-invasive ventilation (NIV) use, and continuous NIV use. Covariates in our models include age at diagnosis, sex, onset location, riluzole use, diagnostic delay, ALSFRS-R scores at the landmark time, and ALSFRS-R rates of change from baseline. Internal and external validation techniques were used.

**Results:** For each of our models and landmark times, we present risk prediction intervals for random sets of patient characteristics. We demonstrate our models’ application for an individual’s personal predicted time-to-event. Our internal and external validation metrics indicate good concordance and overall performance. The time to loss of speech models perform the best for each metric in terms of both internal and external validation.

**Discussion:** Landmarking is an efficient, individualized risk prediction model that is intuitive for both clinicians and patients. Importantly, landmarking can be used for clinical trial modeling, personal planning, and development of real-world evidence of the impacts of treatment interventions.

## Background and Introduction

Amyotrophic lateral sclerosis (ALS) is characterized by progressive disability due to motor neuron degeneration. Its clinical manifestations and functional consequences however are quite varied, for several reasons. First, weakness can begin in any region of the body, including bulbar, limb, or ventilatory muscles, and usually spreads regionally. Second, the relative involvement of cortical vs. spinal motor neurons varies considerably among individuals, such that some suffer principally from spasticity and incoordination of the affected muscles, while others suffer principally from weakness. Finally, the rate of disease progression varies considerably among individuals.

For these reasons, while all people living with ALS develop considerable disability, their individual needs, the timing of those needs, and their survival are quite variable. This variability complicates both clinical research and care. Regarding clinical research, clinical trial development for a highly heterogeneous population, with great variance in expected outcomes and poor predictors of outcome, requires a larger sample size than for populations with a homogeneous and predictable course. Regarding clinical care, individual planning is difficult if one cannot accurately anticipate the development of new medical needs.

While it is not possible to alter the heterogeneity of ALS, it is possible to develop models of progression that can aid in clinical trial design and personal planning. At present, predictive survival models have been developed (1,2). While of some utility, these do not aid in predicting time to other recognized disease state milestones. In this study, we use Landmark Time-to-Event modeling (Landmarking) to demonstrate prediction models for several intermediate times to events that can be used for clinical trial modeling, personal planning, and development of real-world evidence of the impacts of treatment interventions.

Landmarking is a well-established method for dynamic prediction modeling in a variety of fields of study (3–7). Recently, landmarking was used to study survival among those with ALS (2). However, the utility of this approach is not limited to survival as the only primary outcome. As such, we present here a novel application of a well-known dynamic prediction modeling process to predict other ALS milestones. Additionally, we have the benefit of access to an external validation dataset with which to validate our dynamic models.

Overall, landmarking provides a method to compare the changing effects of different covariates, biomarkers, and other prognostic factors as a condition progresses over time. It is an efficient individualized risk prediction model that is intuitive for researchers, clinicians, and patients. Landmarking allows patients to understand how their risk of progression changes in real-time with their unique profile of characteristics. It can be used for clinical trial modeling and design and for analysis of the impact of changes in care as real-world evidence.

## Methods

### Database and Data collection

This study utilizes the Clinic-based Multicenter ALS Natural History Data Collection Study (8), a dataset being compiled by the ALS Natural History Consortium (NHC) which provides clinic-derived information from enrolled participants with a diagnosis of ALS who receive their care from ALS multidisciplinary clinics. Data collection has been ongoing since 2015 (8). The project is deliberately inclusive to be as representative as possible of all people obtaining their care in the participating ALS multidisciplinary clinics. The initial sample used for landmark model development included data from 1977 participants from eight participating ALS clinics in the US. After screening the data (see **Figure 2**), our final sample was N=1557. The sample size for each outcome and each model differs slightly based on timing of events.

All participants provided written informed consent to be included in the database. Current IRB numbers: University of Minnesota (UMN) sIRB: 00019204; Istituti Clinici Scientifici Maugeri: 2687CE; and Centro Clinico NeMO: 247-052017. Prior IRB numbers: UMN: 1501M61381; Northwestern: STU00209860; Saint Louis University: 28018; Temple: 25661; Virginia Commonwealth University: HM20009645; University of Florida: IRB201701910; Henry Ford: 10105; and Providence: 16-123A. Data from Emory University collected through the Clinical Research in ALS Study (CRiALS), IRB: 00078771.

### Data Availability Statement

Data are available upon reasonable request for research purposes. If interested, please reach out to the senior author, Dr. David Walk, for further instructions and availability questions.

### Statistical Models

Landmarking (9–11) was implemented to determine the effect of patient characteristics on disease progression. Landmarking incorporates longitudinal changes in time-to-event modeling to provide updated predictions at pre-specified times after diagnosis, referred to as landmark times. Landmark models allow for flexibility in understanding the effect of various covariates as time progresses and can accurately model time-varying effects of patient characteristics or time-varying covariates such as biomarkers. Using landmark models, a time-to-event model is fit at each prespecified landmark time with an updated, representative risk set of those who have yet to be censored or experience an event at that point in time after diagnosis.

Five outcomes in ALS disease progression were considered: loss of ambulation, loss of speech, gastrostomy, NIV use, and continuous NIV use. We modeled our outcomes using longitudinal assessments of the ALS Functional Rating Scale - Revised (ALSFRS-R) (12). The landmarking process is detailed in **Figure 1**.

**Figure 1:**
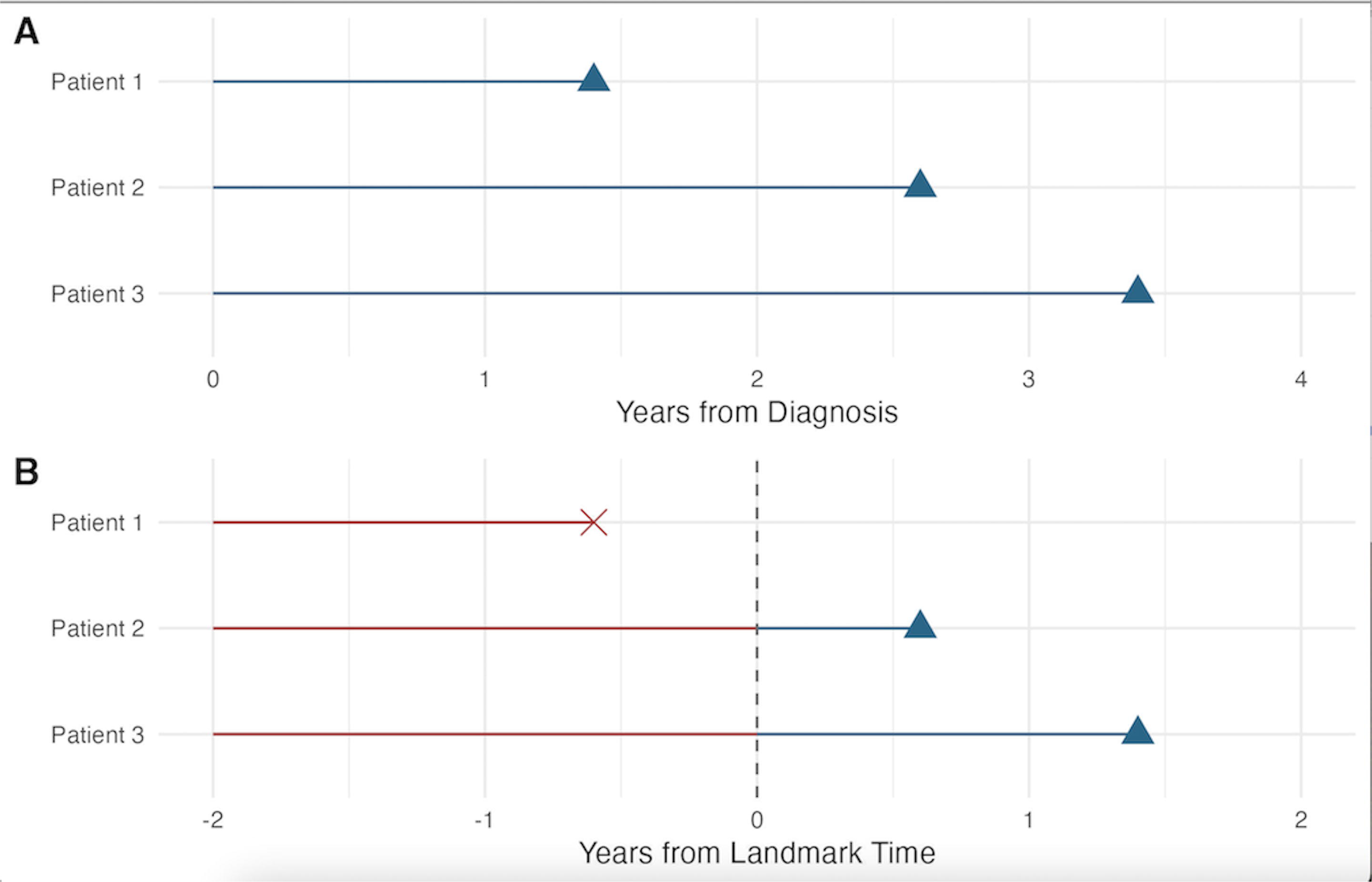
The landmark process is illustrated with a set of three patients with follow up times represented by triangles or Xs. Figure A shows the original follow up time for three patients from diagnosis. All three patients are included in the original model. Figure B shows a landmark model that starts two years from diagnosis. Patient 2 and 3 have yet to experience the event, so they are included in the model (blue portion of line), whereas Patient 1 already experienced the event (designated with an X) and is therefore censored.

Four different landmark times per outcome were applied in this study: six months, one year, two years, and three years past diagnosis. For each participant, the time of event was defined as the first instance of an ALSFRS-R score indicative of each outcome other than gastrostomy. Loss of ambulation was defined as ALSFRS-R question 8 ≤ 1 (Non-ambulatory Functional Movement), loss of useful speech was defined as ALSFRS-R question 1 = 0 (Loss of Useful Speech), NIV usage was defined as ALSFRS-R question 12 ≤ 3 (Intermittent use of BiPAP), and continuous NIV usage was defined as ALSFRS-R question 12 ≤ 1. For gastrostomy, the procedure date was used as the event time. Participants that did not experience an event were considered to be right-censored at their last recorded visit time.

The following covariates were included in each of our models: all four ALSFRS-R subscores (Speech/Swallowing, Fine Motor Skills, Gross Motor Skills, and Respiratory Function), onset location (bulbar vs. limb), age at diagnosis, biological sex, diagnostic delay (time from reported onset of symptoms to clinical diagnosis), time between Last Observation Carried Forward (LOCF) and landmark time, and rate of change for each ALSFRS-R subscore since diagnosis. Edaravone usage was not included in our models as it was not recorded in our validation dataset.

For each landmark time *s*, we implemented a LOCF approach for longitudinal predictors. Each covariate was included in all models. The most recent ALSFRS-R scores were taken to be the representative values of each patient’s ALS severity at the landmark time. We included a model covariate to account for the variability in the length of time passed between landmark time *s* and the LOCF.

### Prediction

For each outcome and each landmark model, we grouped individuals in the risk set into quintiles based upon the model’s linear predictor for rates of progression: very slow, slow, intermediate, fast, and very fast. To visualize our results, we randomly selected three participants per risk group and plotted predicted time-to-event probabilities to compare the trajectories across groups. Furthermore, we present predicted time-to-event curves for typical individuals with a given set of characteristics to illustrate the dynamic prediction modeling process of landmark time-to-event models.

### Validation

For each model, we performed both internal and external validation to determine the overall generalizability of our models. For external validation we used an ALS cohort dataset from Emory University (‘Emory ALS dataset’). The Emory cohort demographic information and a comparison with the NHC dataset can be found in Supplementary Materials.

For internal validation, we calculated a cross-validated concordance value C index and integrated Brier Score (iBS) (13). C index values near 1, perfect predictive ability, indicate good model performance, and values near 0.5, essentially random chance, indicate poor model performance. To calculate our C index values, we implemented five-fold cross validation techniques using the ‘rms’ R package (14).

The iBS, a measure of overall performance and calibration, is a measure of how well our predicted time-to-event risks match up with the observed risks. Values closer to zero indicate better overall performance across the prediction time. For our iBS, we used five-fold cross validation techniques using the riskRegression package (15). All internal discrimination and overall performance metrics are computed with cross-validation techniques, and external validation metrics were computed using the Emory ALS dataset. All performance metrics were evaluated 5 years out from the given landmark time. Analysis was carried out with R version 4.4.0 (16).

## Results

The baseline demographics of the entire cohort of patients with any number of ALSFRS-R visits completed are presented in **Table 1**. Of the total sample, 62.7% were prevalent cases, defined as having been enrolled more than 90 days after date of diagnosis. 978/1977 underwent at least one form genetic testing; of these, 76/572 (13%) had a C9orf72 repeat expansion, 36/475 (8%) had a pathogenic variant in Superoxide dismuatase 1 (SOD1), and 9/449 (2%) had a pathogenic variant in Fused-in-Sarcoma (FUS). Our flow diagram can be found in **Figure 2**. We further filtered the patients from our total 1557 patients for each of the landmark times such that as the landmark time increases, our sample sizes for the landmark models decrease due to events or censoring. For each of the outcomes considered, our 3-year landmark model sample size is between 100 and 200 participants.

**Table 1:**
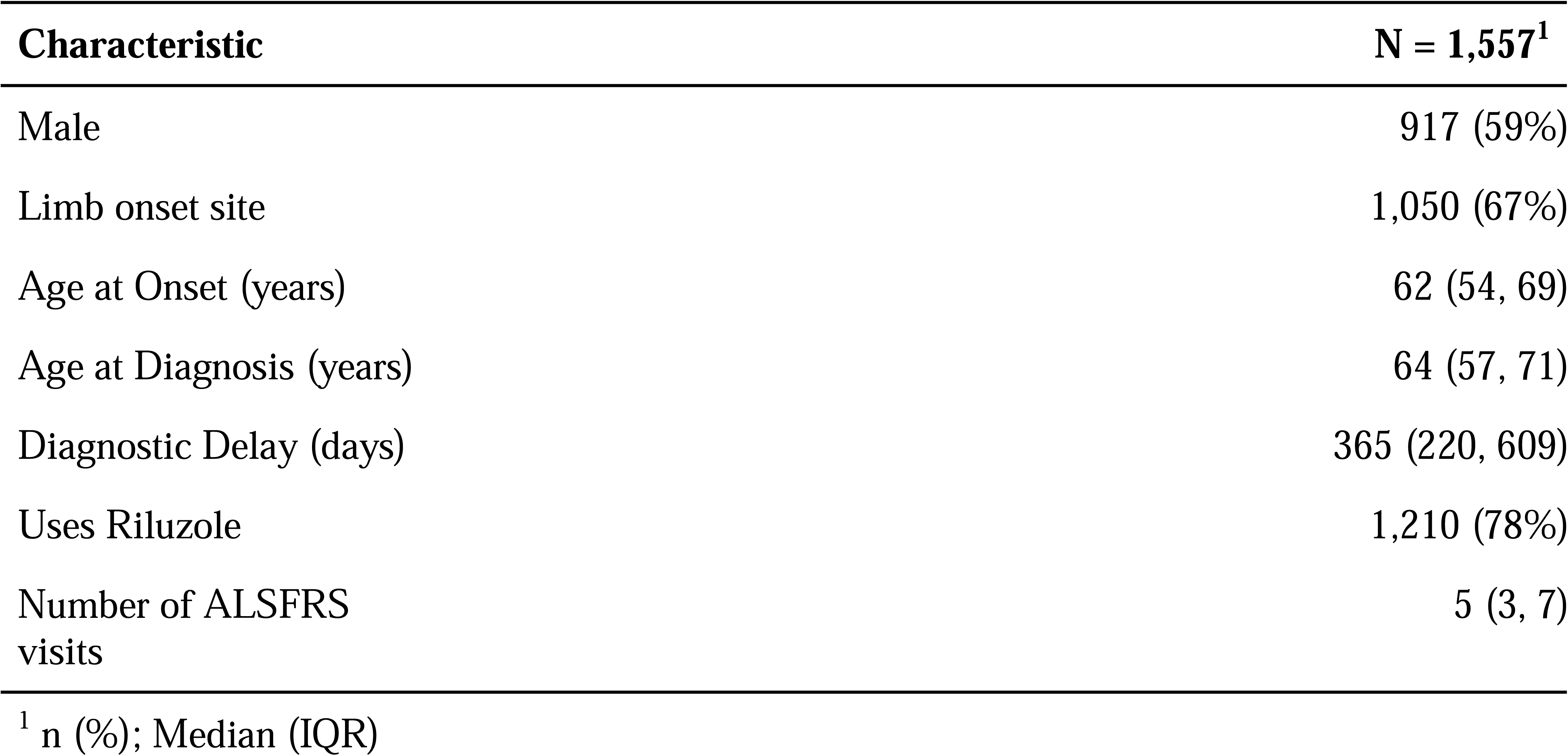
Baseline demographics and medical history for our overall cohort of patients incorporated in this study.

| Characteristic | N = 1,557<sup>1</sup> |
| --- | --- |
| Male | 917 (59%) |
| Limb onset site | 1,050 (67%) |
| Age at Onset (years) | 62 (54, 69) |
| Age at Diagnosis (years) | 64 (57, 71) |
| Diagnostic Delay (days) | 365 (220, 609) |
| Uses Riluzole | 1,210 (78%) |
| Number of ALSFRS visits | 5 (3, 7) |

<sup>1</sup> n (%); Median (IQR)

**Figure 2:**
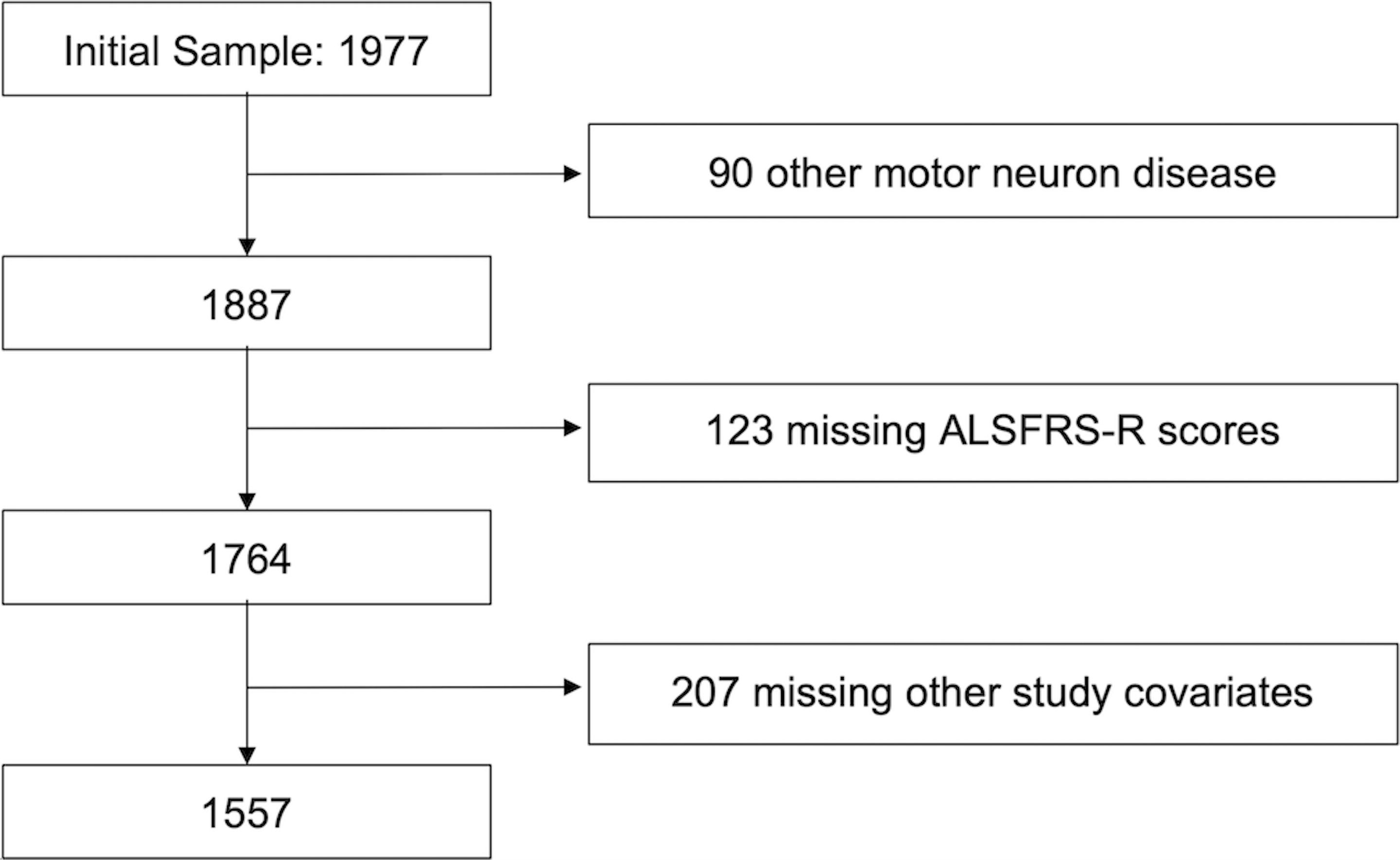
Eligibility criteria used to filter the total database to the sample used in our models. Our total number of subjects after filtering was 1557.

**Figure 3** shows a forest plot of some of the key variables included in our time-to-gastrostomy models. Estimates to the right of the vertical line represent risk factors, while estimates to the left indicate protective factors. For example, higher ALSFRS-R subscores mean less risk. Estimates typically stay consistent among the different landmark models; however, there tends to be a slight bias towards variables having no effect with the later landmark times due to decreased sample sizes later in follow-up. In general, ALSFRS-R subscores, onset location, and diagnostic delays are strongly associated variables across outcome and landmark models.

**Figure 3:**
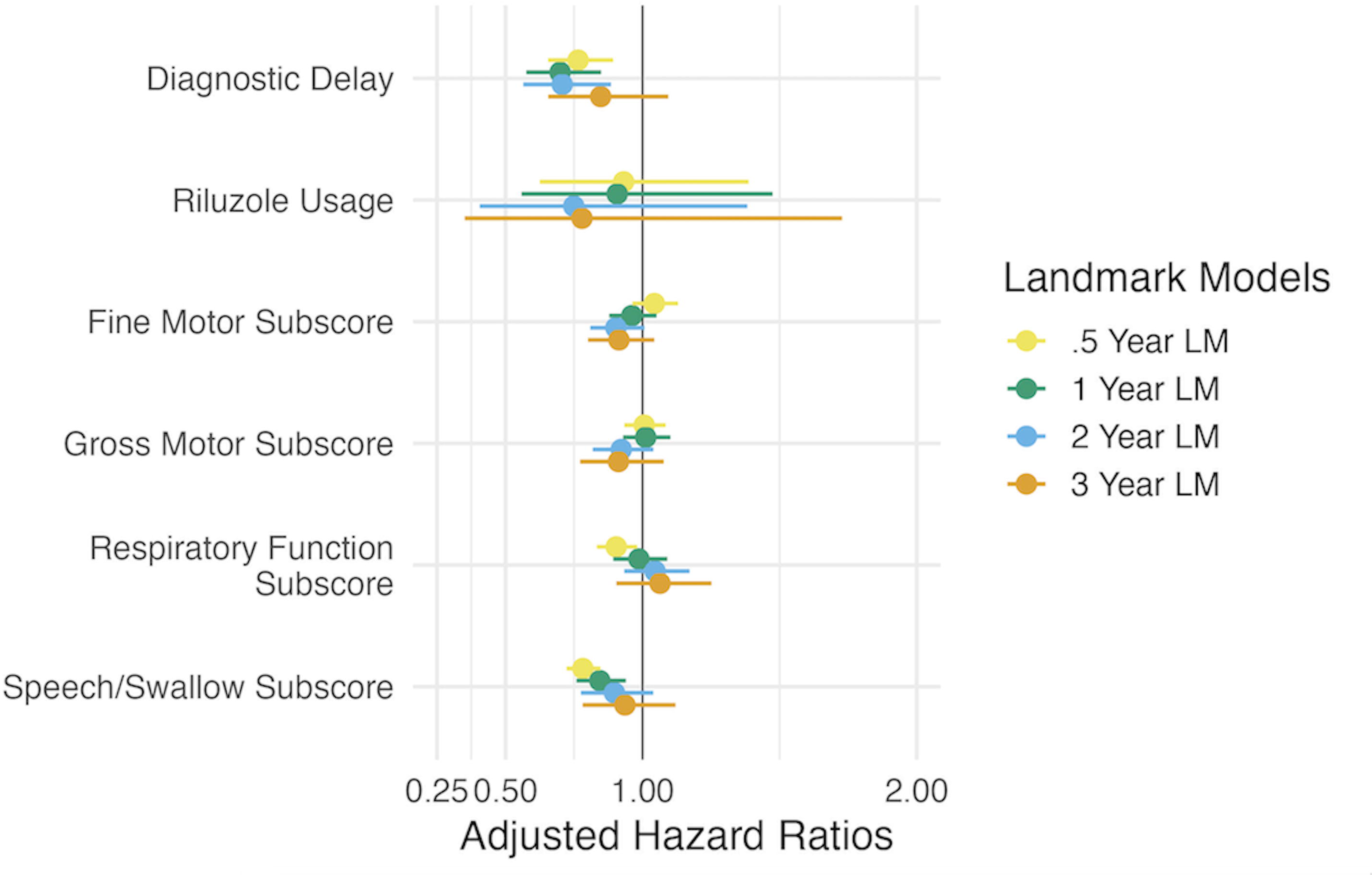
Forest plot illustrating the time-to-event model regression coefficients for four separate landmark models for time to gastrostomy. The dot represents the coefficient estimate and the line intersecting the dot is the 95% CI for the regression estimate. Estimates to the left of the vertical black line represent protective variables and estimates to the right indicate the variable is associated with accelerated progression.

To characterize the clinical application of our models to individual patients, we present our time-to-event prediction curves by classified progression rate for loss of ambulation at the one-year landmark time in **Figure 4**. Each boxplot in this figure contains the predicted survival probabilities of a randomly selected individual patient from within our dataset. Collectively, these lines indicate the ability of our models to separate patients into distinct progression rate groups. Each individual boxplot can be represented further by an individualized predicted time-to-event probability curve as in **Figure 5**.

**Figure 4:**
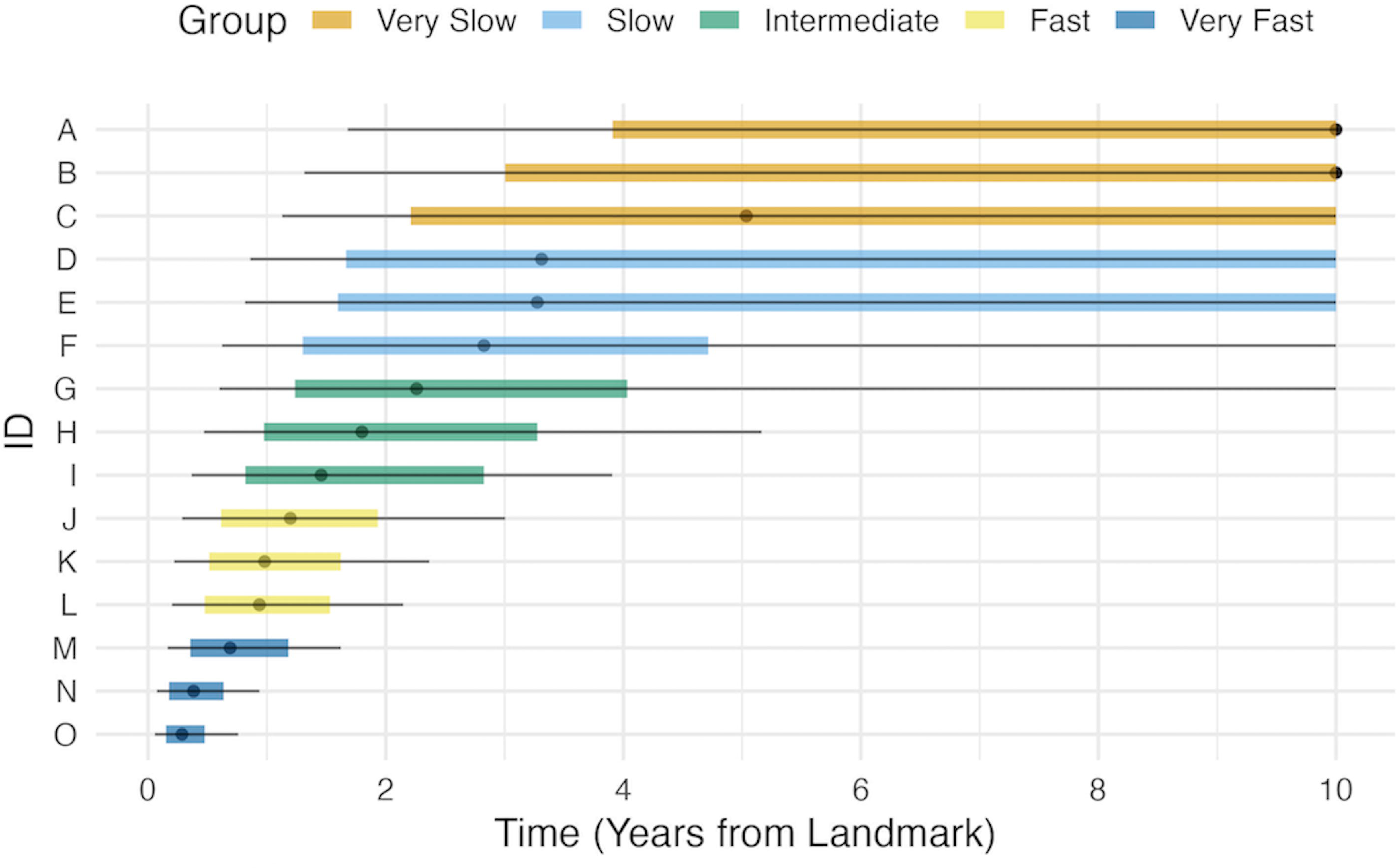
Time-to-event probability predictions at the 1-year landmark time for loss of ambulation, grouped by the Cox proportional hazards model linear predictor (very slow to very fast). Each row represents a summary of an individual’s predicted time-to-event probability curves, given that they had not experienced the event at the 1-year landmark time. The gray dot in the center of each line represents the time at which the predicted probability of not experiencing an event is equal to 50%. The colored boxes represent the times at which the predicted probability of not experiencing an event is between 75% and 25%, and the gray line that intersects the colored boxes for each patient represent the predicted probabilities of 90-10%.

**Figure 5:**
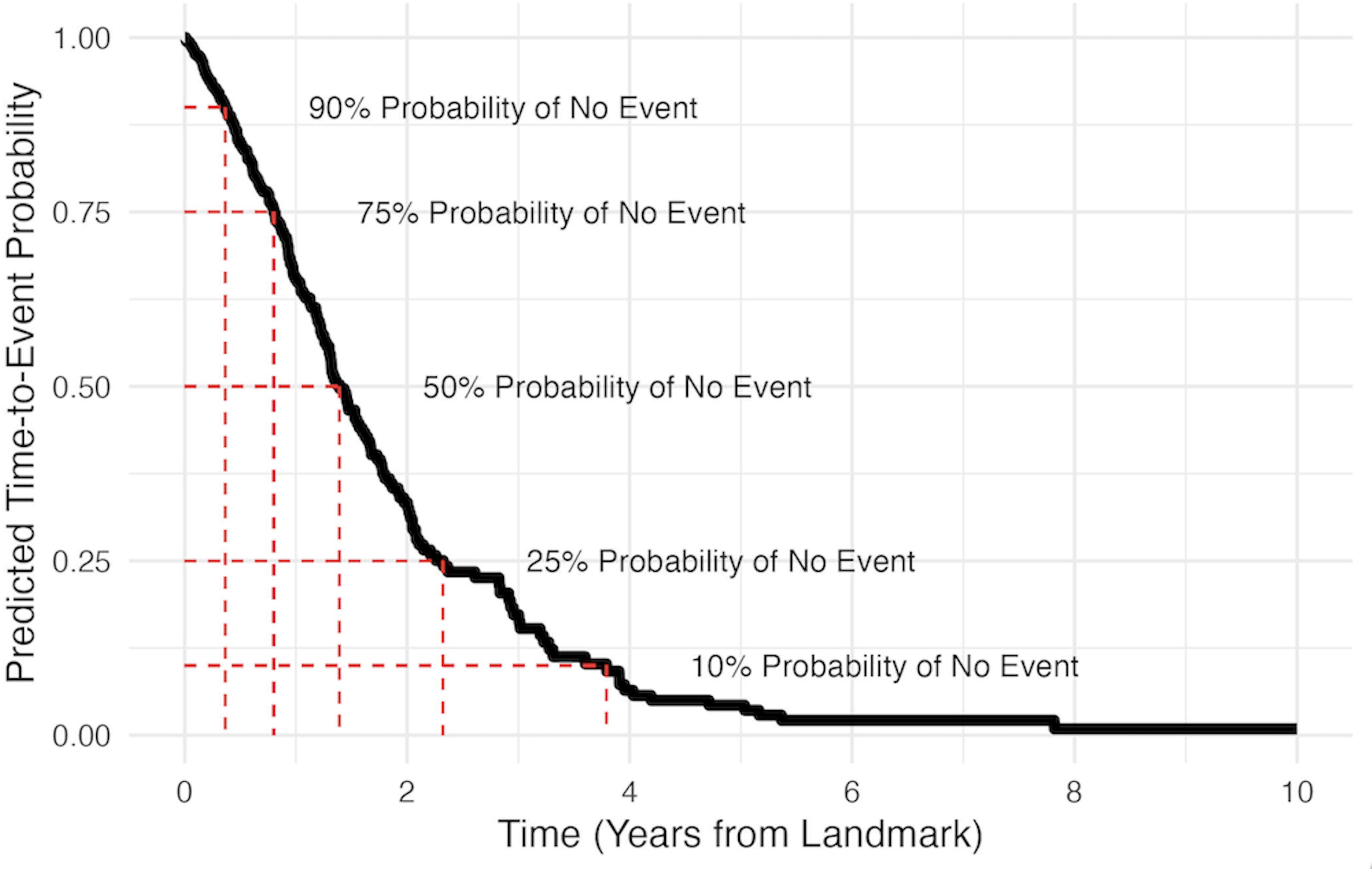
Illustration of an individual time-to-event probability prediction at 1-year landmark time for loss of ambulation. At any given landmark time, we can input a set of characteristics to calculate an individual’s predicted time-to-event curve.

### Performance metrics

We present our discrimination and overall performance metrics in **Table 2**. Across all outcomes, the internally validated C index indicates overall good discriminatory ability, with our time to loss of useful speech and continuous NIV usage models performing the best. Each of the models performed well when applied to the external validation dataset, and in some cases outperformed the internal validation metric. All the discrimination metrics presented a downward trend as the landmark time increases, which reflects loss of power with decreasing sample size as landmark time increases.

**Table 2.** Internally cross-validated and externally validated C index values and iBS from landmark models for each outcome. C index values closer to one and iBS values closer to zero indicate better performance.

|  | Validation Type<sup>a</sup> | 0.5 | 1 | 2 | 3 |
| --- | --- | --- | --- | --- | --- |
| **Loss of Ambulation - C Index** | Internal | 0.775 | 0.793 | 0.750 | 0.699 |
|  | External | 0.752 | 0.740 | 0.713 | 0.661 |
| **Loss of Ambulation - iBS** | Internal | 0.155 | 0.156 | 0.165 | 0.215 |
|  | External | 0.174 | 0.185 | 0.187 | 0.208 |
| **Gastrostomy - C Index** | Internal | 0.745 | 0.669 | 0.673 | 0.610 |
|  | External | 0.782 | 0.803 | 0.736 | 0.759 |
| **Gastrostomy - iBS** | Internal | 0.170 | 0.174 | 0.177 | 0.198 |
|  | External | 0.164 | 0.158 | 0.159 | 0.128 |
| **NIV - C Index** | Internal | 0.683 | 0.720 | 0.616 | 0.557 |
|  | External | 0.666 | 0.662 | 0.648 | 0.721 |
| **NIV - iBS** | Internal | 0.179 | 0.173 | 0.200 | 0.259 |
|  | External | 0.172 | 0.183 | 0.189 | 0.204 |
| **Continuous NIV - C Index** | Internal | 0.861 | 0.852 | 0.854 | 0.827 |
|  | External | 0.703 | 0.689 | 0.756 | 0.777 |
| **Continuous NIV - iBS** | Internal | 0.075 | 0.082 | 0.087 | 0.094 |
|  | External | 0.144 | 0.137 | 0.097 | 0.072 |
| **Loss of Useful Speech - C Index** | Internal | 0.870 | 0.884 | 0.863 | 0.785 |
|  | External | 0.843 | 0.851 | 0.816 | 0.808 |
| **Loss of Useful Speech - iBS** | Internal | 0.103 | 0.099 | 0.096 | 0.140 |
|  | External | 0.133 | 0.116 | 0.116 | 0.110 |

Our iBS values for the internally validated models indicate good performance between the landmark time and the five years following. Each outcome’s iBS values tended to increase as the landmark time increased in the internal validation models, while the external validation iBS values for time to gastrostomy and time to loss of useful speech tended to decrease slightly. The loss of useful speech and continuous NIV usage models had the best overall performance among all the outcomes considered. Again, the internally validated and externally validated iBS values were similar, indicating good generalizability to the Emory ALS dataset. In most cases the internal metrics were better than the external metrics because the model was built with the ALS NHC dataset. However, time to gastrostomy models performed better in the Emory ALS dataset than in the NHC dataset.

### Rshiny Web Application

An interactive web application is being developed using Shiny (17) to demonstrate the utility and flexibility of the landmarking time-to-event modeling approach. Contained within this web application are group-based time-to-event probability prediction results for each outcome and for each landmark model time considered.

Similarly, for each outcome and for each landmark time, we offer the ability to view individualized time-to-event prediction curves for a given set of characteristics that are included in the models. A clinician can use this web application to obtain an estimated time-to-event prediction probability for any of the considered intermediate outcomes in ALS progression.

## Discussion

We implemented landmark time-to-event analysis for five intermediate outcomes in ALS and demonstrated the dynamic modeling approach from date of diagnosis. We modeled our time-to-event analysis using longitudinal records from the ALS NHC, which represents a broad clinic-based population of people living with ALS. For each outcome and landmark time, we presented prediction diagnostic statistics, provide measures of internal and external model validation, and demonstrated the application of our prediction models for patients and clinicians via our group and individual prediction plots.

Our models contribute to time-to-event modeling of clinically meaningful intermediate events in ALS. Previous studies with natural history datasets have developed personalized prediction models (1,18), and the landmark time-to-event approach adds a dynamic component to the modeling process. Westeneng et. al, for example, used a rate of change for ALSFRS-R scores across all of follow-up rather than modeling with visit-specific rates of change. Furthermore, many studies (1,2,18,19) that seek to develop personalized prediction models for ALS looked at survival only or composite endpoints for death, tracheostomy, or use of NIV for more than 23 hours per day. Our models look at intermediate events in the ALS progression that can be used in clinical trials modeling, assessment of real-world evidence, and personal planning. Though we focused solely on intermediate outcomes, the landmark approach can be extended to survival or time-to-event composite endpoint modeling as well. This approach can also be utilized for time to advancement in ALS staging, such as with King’s or MiToS (20,21).

Our models performed well in both prediction metrics used. Our early landmark models consistently had a C-index value greater than 0.7, with some outcomes such as loss of useful speech and continuous NIV usage yielding similar values in the later landmark models as well. Furthermore, we demonstrate that our models generalize to the Emory ALS dataset via our external validation iBS and C Index values. As our database continues to grow, we will improve our models’ predictive capability.

### Limitations

The inclusion of prevalent cases into our study likely biased our results and predictions toward slower-progressing individuals. Furthermore, because ALS has a short median survival time and these outcomes progress quickly, we lose power as landmark times increase, which diminishes our predictive ability in later landmark models.

Because we used a real-world dataset, the frequency of visits for individuals varies considerably. We have ALSFRS-R scores at most every three months, and as such there are gaps in our understanding of the progression of disease. Furthermore, we defined most of our outcomes based upon ALSFRS-R scores, so we had to model with the assumption that many of these events of interest did not happen exactly on the day the data were recorded. In addition, we were restricted to ALSFRS-R being our only time-varying, longitudinal covariate.

As the number of treatments for ALS grows, the natural history will change. In addition, therapies specific to individual characteristics, such as antisense oligonucleotides for specific pathogenic variants, will need to be included in modeling. Furthermore, edaravone was not included in our models as its usage was not included in our validation dataset.

Finally, we are only modeling one event path at a time with this landmarking approach, and therefore do not explicitly include the effect of other intermediate outcomes on risk of intermediate outcome. If the reader is interested in a semi-competing risks modeling approach for ALS time-to-event analyses, we refer you to our companion paper, Arguedas et al., ‘Risk Prediction for ALS using Semi-competing Risks Models with Applications to the ALS Natural History Consortium Dataset.’

### Next steps

As genetic testing becomes the standard of care in ALS, we will add genotype to phenotype information in our list of baseline characteristics. In addition, we will incorporate additional clinical measures and blood biomarkers into our model and determine the degree to which they impact model performance. Finally, we plan to model the applicability of our landmarking model as a tool for propensity-score matching and pragmatic clinical trials.

## Data Availability

Data are available upon reasonable request for research purposes. If interested please reach out to the senior author Dr. David Walk for further instructions and availability questions.

## Acknowledgement

This project was supported by the FDA’s Office of Orphan Products Development under grant number FD-R01-0007630. Its contents are solely the responsibility of the authors and do not necessarily represent the official views of the FDA nor FDA’s Office of Orphan Products Development.

## Supplementary Materials

**Table S.1** compares the NHC sample used to build our models and the Emory dataset used to validate them. The sample size from Emory reflects the eligibility criteria used for the NHC sample. Contained within the table are both the p value for the difference in sample characteristics (via Pearson’s Chi-Squared tests) and the standardized mean difference (SMD) [1]. The SMD is an effect size metric used to quantify the amount by which variables differ between two groups.

Established criteria for interpreting SMD values are as follows: a SMD of 0 indicates that the groups are identical, a value below 0.1 is considered inconsequential, values between 0.1 and 0.2 are small differences, and anything about 0.2 is considered large.

There were negligible differences in the proportion of males in the two samples, but small differences existed in the proportion of participants with limb onset, age of onset, and age of diagnosis. Large differences were observed in the number of visits for the two samples and the proportion of participants that have taken Riluzole.

**Table S.1:** Presented here are the Emory dataset demographics and baseline characteristics in comparison to the NHC dataset. The Emory dataset was used as a validation set for our landmark models.

| Characteristic | Emory (N = 1,497)<sup>1</sup> | NHC (N = 1,507)<sup>1</sup> | SMD<sup>2</sup> | p-value<sup>3</sup> |
| --- | --- | --- | --- | --- |
| Male | 877 (59%) | 917 (59%) | 0.006 | 0.89 |
| Limb onset site | 1,090 (73%) | 1,050 (67%) | 0.12 | 0.001 |
| Age at Onset (years) | 61 (52, 69) | 62 (54, 69) | 0.12 | .002 |
| Age at Diagnosis (years) | 62 (53, 70) | 64 (57, 71) | 0.16 | <0.001 |
| Uses Riluzole | 491 (33%) | 1,210 (78%) | 1.01 | <0.001 |
| Number of ALSFRS visits | 3.00 (2.00, 5.00) | 4.00 (3.00, 7.00) | 0.29 | <0.001 |

<sup>1</sup> n (%); Median (IQR)
<sup>2</sup> Standardized Mean Difference
<sup>3</sup> Pearson's Chi-squared test

